# “They could just come, they peep, then they get back”: COVID-19 patients’ experiences of care in isolation in Nairobi, Kenya

**DOI:** 10.64898/2026.10.20.26366069

**Authors:** Jeremiah M. Nganda

## Abstract

Isolation is a core measure of outbreak control, yet it reduces patients’ contact with health workers and removes the support of family. During the COVID-19 pandemic, health workers became the main human contact for isolated patients while themselves contending with fear of infection and stigma. Little is known about how patients in sub-Saharan Africa experienced health workers’ conduct under these conditions. This study explored how adults isolated with COVID-19 in Nairobi, Kenya, experienced interactions with health workers. I conducted a qualitative descriptive study using in-depth interviews with 15 adults who had recovered from COVID-19 after hospital isolation in Nairobi between 2020 and 2022. Participants were recruited through purposive and snowball sampling and interviewed in February and March 2023. Data were analysed using reflexive thematic analysis, and the resulting codes were mapped onto the Bohren et al. typology of mistreatment. Four themes were developed. First, some health workers offered attentive care, giving patients hope through encouragement and reassurance. Second, contact was brief and distant, a pattern that health workers themselves attributed to fear of infection. Third, care was organised around routine monitoring, so that deterioration and emotional distress often went unnoticed. Fourth, some patients encountered ridicule of their distress and perceived that patients were treated unequally. Most experiences corresponded to existing domains of the typology but distancing driven by fear of infection did not. Isolation increased patients’ dependence on health workers at the same time as it created conditions that drew health workers away from them. Fear-driven distancing emerged as an additional domain of mistreatment in outbreak settings. Unaddressed, these patterns may compromise patient safety and weaken the trust on which outbreak responses depend. Preparedness plans should set minimum contact standards for isolated patients, include psychosocial assessment in routine monitoring and address health workers’ fear of infection.

## Introduction

Isolation is among the oldest and most widely used measures of outbreak control, but it carries a well-documented cost for the people isolated. Studies have reported that isolated patients reported more anxiety and depression, had fewer contacts with health workers, and experienced more adverse events linked to failures of supportive care than other patients [1,2]. In addition, a study reported that attending physicians examined 35% of patients in contact isolation compared with 73% of patients who were not isolated [3]. Isolation therefore tends to reduce the human contact that patients most need when they are frightened and unwell.

The COVID-19 pandemic magnified this problem. Visiting restrictions removed relatives, who ordinarily provide comfort, advocacy and practical help, and were associated with loneliness, lower psychosocial wellbeing and agitation among patients [4]. Health workers became the main, and often the only, human contact for isolated patients. Patients in some instances described relying on nurses for companionship and reassurance, communicated through personal protective equipment (PPE) that obscured faces and limited touch [5,6]. While the focus has been on the patients, the health workers, seem to have carried a heavy burden. Studies have estimated that more than one in five experienced anxiety or depression early in the pandemic [7], and other studies have described exhaustion, uncomfortable PPE, scarce resources and fear of infection [8]. Health workers who were themselves isolated with COVID-19 reported stigma from staff at isolation centres and called for more compassionate care [9]. Research on interpersonal interactions during the pandemic has nonetheless focused mainly on aggression by patients and relatives towards health workers [10]. How patients experienced the conduct of health workers has received far less attention.

Yet, the quality of interaction between patients and health workers is a dimension of quality of care in its own right. Mistreatment of patients in health facilities has been studied most extensively in maternity services, where a global systematic review documented physical and verbal abuse, discrimination and neglect across a wide range of countries [11]. In Kenya, about one in five women reported disrespect or abuse during childbirth, most often non-dignified care and neglect [12], and health workers attributed mistreatment to stress, staff shortages, facility culture and bias based on social status [13,14]. Similar patterns of negligence, impoliteness and discrimination have been documented in general hospital care [15]. Disrespect is increasingly understood as a product of social and health system conditions rather than of individual character alone [16].

If such patterns take hold in isolation units, their consequences could extend beyond individual distress. Isolated patients are often acutely ill, so missed monitoring poses a risk to safety [1]. Poor treatment can also deter people from using services[17]. More broadly, everyday encounters with health workers shape public trust in the health system [18], and trust underpins the testing, disclosure and acceptance of isolation on which outbreak responses depend. These concerns are growing more pressing. The World Health Organization reported a 63% increase in zoonotic outbreaks in Africa between 2012 and 2022 compared with the previous decade [19], and isolation is therefore likely to remain a recurring experience for patients in the region.

Evidence from patients in sub-Saharan Africa remains limited. In Ghana, Osei and colleagues found that patients isolated with COVID-19 experienced fear, loneliness and stigma; most praised staff, although some described nurses as distant and disrespectful [20]. The conduct of health workers was, however, a secondary finding of that study. In Kenya, early cases of COVID-19 were admitted to hospitals and isolation facilities. From June 2020, home-based isolation was introduced for mild cases, while patients with more severe illness, or without space to isolate at home, continued to be isolated in health facilities [21]. To our knowledge, no study has examined Kenyan patients’ accounts of how health workers treated them during COVID-19 isolation.

### Theoretical Framework

We used the typology of mistreatment developed by Bohren and colleagues as the conceptual framework for this study [11]. Derived from a mixed-methods systematic review of women’s experiences of facility-based childbirth, the typology organises mistreatment into seven domains: physical abuse; sexual abuse; verbal abuse; stigma and discrimination; failure to meet professional standards of care; poor rapport between patients and providers; and health system conditions and constraints. Two features make it suitable for this study. First, it spans the interpersonal and the structural, placing individual acts such as ridicule alongside conditions such as staff shortages that shape how care is delivered. Second, its domain of poor rapport encompasses ineffective communication, lack of supportive care and loss of autonomy, which allows supportive and unsupportive interactions to be examined within one frame. The typology has been widely used in maternity care research in Kenya and elsewhere [12,13], but it has rarely been applied to other areas of care, and not, to our knowledge, to outbreak settings, where infection control reshapes almost every encounter between patients and health workers.

This study therefore explored how adults isolated with COVID-19 in Nairobi experienced their interactions with health workers. We asked how these experiences corresponded to the domains of the typology, where they departed from it, and what this implies for care in future outbreaks.

## Methods

### Study design

The study employed a qualitative descriptive design with in-depth interviews, which is suited to producing a detailed account of participants’ experiences in their own words. This paper draws on a wider study of lived experiences of patients who recovered from COVID-19 in Nairobi; findings on participants’ use of self-talk as a coping strategy during isolation are reported elsewhere [22]. The present analysis addresses a distinct question, patients’ experiences of health workers’ conduct, and reports different themes and largely different data.

### Setting and participants

The study was conducted in Nairobi County, Kenya. Participants had been admitted with COVID-19 to selected leading hospitals in Nairobi between 2020 and 2022 and had been isolated for varying lengths of time. Eligible participants had been diagnosed with COVID-19 and admitted to one of the selected hospitals, had fully recovered, lived in Nairobi County, were aged over 18 years and could communicate in English. Participants were recruited through purposive sampling, to include people with direct experience of hospital isolation, and snowball sampling, in which early participants introduced other recovered patients. Fifteen participants were interviewed. Preliminary analysis in the wider study indicated that data saturation was reached after the twelfth interview, and all 15 interviews were retained for analysis [22].

### Data collection

Interviews were conducted between February and March 2023. Interviews were conducted in person and lasted between 30 to 60 minutes. Interviews were done in English, but where participants used Kiswahili expressions, these were translated by the research team. Interviews were audio-recorded with participants’ consent and transcribed verbatim by two research assistants.

### Data analysis

The study used reflexive thematic analysis [25,26], which proceeded in two stages. In the first stage, transcripts were read repeatedly and coded inductively, line by line, for any account of contact with health workers, whether positive or negative. Codes were grouped into themes, which were reviewed against the coded extracts and the full dataset and refined through discussion within the research mentors.

In the second stage, we mapped the codes and themes onto the seven domains of the Bohren et al. typology [11]. For each theme, we asked which domain or domains it corresponded to, whether it fitted the domain’s definition, and what in the data the typology did not capture. The typology was used as a sensitising framework rather than a coding frame: themes were developed inductively before the mapping, and data that did not fit any domain were retained and examined rather than set aside. Accounts that ran against emerging patterns were examined as deviant cases.

### Rigour and reflexivity

Credibility was supported by team discussion of codes and themes, an audit trail of analytic decisions, and attention to deviant cases. The author has not worked in the hospital and had no relationship with any of the hospitals where the participants were isolated. Interpretations we checked against participants’ own words and sought neither to excuse nor to condemn individual health workers. Quotations are labelled by participant number to protect the identify of the participants.

### Ethics

The study was approved by the Daystar University Institutional Scientific and Ethical Review Committee, and a research permit was obtained from the Kenya National Commission for Science, Technology and Innovation (NACOSTI/P/23/23220). All participants gave written informed consent, including consent for audio recording. Transcripts were anonymised before analysis.

## Results

### Participants

Fifteen recovered patients took part: nine women and six men, all aged over 18 years. Their occupations included cashier, professional model, businessperson, university student, pharmacist, government agency employee and casual labourer. All had been hospitalised and isolated with COVID-19 in Nairobi between 2020 and 2022.

### Overview of themes

Four themes were developed: attentive care that gave hope; brief and distant contact; routine monitoring and missed distress; and ridicule and unequal treatment. Some participants experienced both supportive and disrespectful encounters within a single stay, sometimes from different health workers responding to the same distress. What participants received appeared to depend less on the facility than on which health worker entered the room and on what that health worker chose to do in the limited time available. The sub-sections below present the results organized according to the themes.

### “They encouraged me”: attentive care that gave hope

Several participants described health workers who encouraged and reassured them and helped them imagine recovery. For patients frightened by their own breathing, hearing that sicker people had recovered was a turning point:

> “I used to doubt… when I was going to sleep, whether I would not wake up because of the breathing… One of the nurses… came and found me when I was awake and asked me why I was not sleeping… I told them that I had fears… but they encouraged me, they told me of how even worse people had come before in the hospital and they had healed.” (P004)

This encounter began with attention rather than with a clinical task. The nurse noticed that the patient was awake at night and asked why, which opened the way for the patient to name a fear of dying in their sleep. The reassurance that followed drew on the nurse’s own experience of the ward: others who had been more severely ill had recovered. In a setting where patients had no family to interpret what was happening to them, health workers were the only people who could offer this kind of comparison, and it carried weight because it came from someone who had seen many patients through the illness.

Other participants recalled encouragement that was personal and repeated, from a nurse who “every time she would come… tell me that you know you look like someone who wins in everything… she really encouraged me a lot” (P002). Health workers also gave patients a sense of time. One participant was told that isolation would last “just like 2 weeks… depending on how… your body is going to react” (P010). For patients confined to a room with no clear end in sight, a bounded period made the experience easier to endure.

The most extended account of support came from a mother who had learnt that her baby had also tested positive. A male health worker found her crying, asked whether she was in pain, asked whether she was willing to talk and, when she said that the matter was personal, persuaded her to speak. He then reframed her distress around her child:

> “He told me… you are here, and your baby is there, the more you cry the more you affect your body… what if you… get soaked in all this pity… and you don’t make it, what will happen to that baby? So he talked to me like I should now try to be positive… I have to do this by getting better, I go back to her.” (P009)

The health worker did not dismiss her distress. He first established what lay behind it and then connected her recovery to the child she wanted to return to, giving her a reason to cope rather than an instruction to stop crying. This exchange shows the sequence that characterised supportive care across the accounts: noticing distress, asking about it, and responding to what the patient said.

These acts were brief, which suited the limited contact that isolation allowed, and they corresponded to the supportive end of the typology’s domain of rapport between patients and providers. None, however, was required by protocol. Each depended on an individual health worker choosing to notice, to ask and to stay a little longer. Supportive care was therefore real, but it was not reliable; whether a patient received it depended on who was on duty.

### “They peep then they get back”: brief and distant contact

Contact with health workers was almost always brief. Participants described health workers who appeared at the door, completed what was needed and left:

> “No, they could just come, they peep, then they get back, they couldn’t stay for long… no one was there to stay for long.” (P008)

The phrase “they peep” conveys how participants experienced these encounters: as glances from the doorway rather than visits to the bedside. The participant’s emphasis that “no one” stayed suggests that this was not the behaviour of one health worker but the general pattern of care in the unit. Participants understood this distance as driven by fear, in part because health workers said so. The same participant recalled one health worker’s explanation:

> “She was telling me that I also fear to be infected, I also am a human being, so not that we are secluding you here that maybe you are a Satan or maybe what, we are just trying to contain the spread of the thing.” (P008)

This explanation acknowledged the health worker’s own vulnerability, presenting her distance as self-protection rather than rejection of the patient. It also addressed a fear the health worker appeared to anticipate - that isolation meant being treated as evil or contaminating (“a Satan”). By describing isolation as a measure against the virus rather than against the person, she turned an experience of exclusion into one the patient could understand and accept. The account shows that the same distance could be experienced as rejection or as reasonable caution, depending on how it was presented.

Distance was also built into how care was organised. One participant noted that only two nurses “were the only ones who were allowed to come and see me in the room,” checking “after some six hours” for temperature and vital signs (P010). Restrictions on who could enter a room, and how often, were part of infection control protocols, and they set the outer limits of contact. Within those limits, individual health workers kept each entry short. The data therefore point to distance operating at two levels: protocols that limited contact by design, and health workers who, fearing infection, minimised the contact that protocols allowed.

*“The only time they checked on me was the first day”: routine monitoring and missed* distress

When contact was limited, it was organised around a narrow routine of vital signs and medication. Participants described what fell outside that routine. One said that health workers checked on them properly only on admission, when they handed over the means of monitoring:

> “You know, surprisingly the only time they checked on me was the first day I came in… because they gave me thermometer for checking my temperature and something for the oxygen and gave me five medication for five days.” (P013)

The word “surprisingly” signals that the participant had expected more. Having been given a thermometer, an oxygen monitoring device and a course of medication, the participant was in effect left to monitor their own condition. This delegation may have been a reasonable way to limit exposure for patients who were stable, but it also shifted to patients the responsibility for recognising their own deterioration. The same participant described patients in acute distress whose needs went unanswered:

> “Most people who were really affected were wailing, I could hear them… the one who I can say I talked to was the one who was gasping for the air but he could not speak properly… but the unfortunate part is that the nurses were not coming very fast to assist.” (P013)

This account describes a ward in which patients could hear one another’s suffering but could not summon help quickly. For a patient struggling to breathe, delay in assistance is a matter of safety, not only of comfort. The participant’s position as a witness also matters. Hearing other patients wail and gasp without a prompt response is likely to have added to the fear that isolated patients already carried.

Emotional and spiritual distress was also missed. One participant, crying and praying aloud, recalled a health worker who noticed but did not stay:

> “I was in deep conversation with myself and God… telling God why me, please heal me… she told me ‘are you okay’ and I… just nodded my head. So tears were falling… I was crying and talking to God.” (P014)

The health worker asked the right question, but a nod from a patient in tears was accepted as an answer. The exchange illustrates how a check on wellbeing can be completed as a task without engaging with what the patient is experiencing. Participants also noted that health workers did not discuss ways of coping with isolation (P015), an observation examined in more detail elsewhere [22].

On the contrary, the participant who reported being checked only on admission nonetheless described being “treated very well” in comparison with others. Being left alone, rather than being treated badly, counted as good care. This suggests that participants’ expectations adjusted to what isolation offered, and that measures of patient satisfaction alone may understate the gaps in care that participants described.

### “Ni kawaida ya COVID patients”: ridicule and unequal treatment

A few participants described responses to their distress that they experienced as mockery. The mother described in Theme 1 had broken down on learning that her baby had tested positive:

> “The day I broke down, when I realised my baby was positive, I cried… and then there were nurses outside and they started laughing, they were like, she will get used to it, *hajakubali ako na COVID, ni kawaida ya COVID patients* [she hasn’t accepted she has COVID, it’s normal for COVID patients].” (P009)
>
> “Yeah, that was very harsh, they didn’t know why I cried, maybe I was in a lot of pain, they didn’t come to check what’s going on.” (P009)

The nurses’ remark placed her tears in a familiar category: a “COVID patient” who had not yet accepted the diagnosis. Within that category, her distress was predictable and therefore did not require a response. The real reason for her tears, her baby, never surfaced, and her second statement makes clear that what hurt was not only the laughter but the failure to ask. The contrast with the health worker in Theme 1, who asked why she was crying and responded to her answer, shows what the category concealed. Seen together, the two encounters suggest that the difference between ridicule and support lay less in the time available than in whether the health worker treated the patient as an individual or as a type. Participants also perceived that patients were treated unequally:

> “For some people who got it [they] were being treated badly… for me I am a tough nut to crack… I was treated very well, not the way people were sidelined… those who were sidelined, it was really a bad experience for them.” (P013)

This participant attributed their own good treatment to their demeanour (“a tough nut to crack”). The account suggests that patients who appeared composed and undemanding may have fared better than those who were visibly distressed. If so, the patients who most needed attention may have been the most likely to be sidelined, a pattern that would compound the vulnerability of those already struggling to cope with isolation.

### Mapping the themes onto the typology of mistreatment

Table 1 summarises how the four themes correspond to the domains of the typology. Physical and sexual abuse were not reported. Verbal abuse, stigma and discrimination, failure to meet professional standards of care, poor rapport and health system constraints were all represented. Supportive care appeared as the positive pole of the rapport domain. One pattern did not fit any domain: distancing that health workers attributed to their own fear of infection. It was neither neglect arising from indifference nor solely a consequence of resource constraints, but a self-protective response to a real threat.

**Table 1.** Themes mapped onto the domains of the Bohren et al. typology of mistreatment [11].

| Theme | Corresponding domain(s)<br>of the typology | Illustrative evidence |
| --- | --- | --- |
| 1. Attentive care that gave hope | Poor rapport between patients and providers (positive: supportive care) | A nurse noticing a sleepless patient and offering hope (P004); a health worker drawing out a mother's distress (P009) |
| 2. Brief and distant contact | Health system conditions and constraints.<br>Proposed additional domain: fear-driven distancing | "I also fear to be infected, I also am a human being" (P008); two nurses permitted, checks about every six hours (P010) |
| 3. Routine monitoring and missed distress | Failure to meet professional standards of care (neglect); poor rapport between patients and providers | Monitoring only on admission and slow response to a breathless patient (P013); a nod accepted as an answer (P014) |
| 4. Ridicule and unequal treatment | Verbal abuse; stigma and discrimination | Laughter at a crying mother, normalised as normal for such patients (P009); patients "sidelined" (P013) |
| 5. Not reported | Physical abuse; sexual abuse | — |

## Discussion

This study found that COVID-19 isolation created a paradox of care. By removing families, isolation made health workers patients’ only human contact and raised the significance of every encounter. At the same time, the conditions of isolation, including the threat of infection, scarce staff and PPE, and protocols that restricted contact, drew health workers away from patients. Participants experienced the result in both directions. A health worker who noticed a sleepless patient and spoke of others who had recovered could change how that patient faced the night. A health worker who looked in and left, accepted a crying patient’s nod, or laughed at a mother’s tears deepened the isolation that patients were already enduring.

The patterns participants described were shaped by the conditions in which health workers worked. The Bohren et al’s typology itself recognises that mistreatment arises not only from individual behaviour but from health system conditions and constraints [11], and our findings support that view. Before the pandemic, isolated patients in high-income settings received half as many contacts per hour from health workers as other patients [2], and attending physicians examined them half as often [3]. Distance is therefore partly a structural feature of isolation, which a novel and frightening pathogen intensified. During COVID-19, health workers in many settings reported fear of infection, exhaustion and shortages of PPE [7,8], and health workers who were themselves patients reported stigma from isolation-centre staff [9]. In Kenyan maternity care, providers attributed mistreatment to stress, staff shortages and facility culture [13]. This evidence suggests that the brief contact, routine monitoring and occasional ridicule described by our participants were, at least in part, predictable responses to the conditions of isolation work rather than simply the failings of individuals.

Structural explanations do not account for everything, however. Within the same wards and under the same constraints, some health workers noticed, asked and encouraged, while others did not. One participant in this study, a mother, received both ridicule and support during a single stay. That supportive care depended on individual choice, rather than on how care was organised, is an important finding. It means that good relational care in isolation was available, but largely by chance.

Participants’ experiences corresponded to the domains of the Bohren et al’s typology. Ridicule, neglect and slow responses mirror findings from Kenyan maternity services, where non-dignified care and neglect were the most commonly reported forms of mistreatment [12], and from general hospital care in Indonesia, where both patients and nurses identified negligence, impoliteness and dismissal, and patients in particular emphasised discrimination [15]. Participants’ perception that some patients were “sidelined” echoes Kenyan providers’ accounts of preferential treatment based on social status [13]. These parallels suggest that the typology applies beyond maternity care and can provide a common language for examining patients’ experiences across services.

The typology assumes that the health worker is present and is concerned with what happens during contact. In isolation, health workers’ own fear limited their presence. Participants did not describe this distance as cruelty; they described health workers protecting themselves, and in one case a health worker said so directly. The fear was real and widely reported during COVID-19 [7,8]. This suggests that fear-driven distancing is an additional domain of mistreatment for outbreak settings. It differs from neglect because it arises from a reasonable concern for one’s own safety rather than from indifference, and from health system constraints because it can operate even where protocols would permit more contact. The two (fear-driven distancing and neglect) differ greatly. Neglect calls for supervision and accountability, whereas fear-driven distancing calls for protection, reassurance and practical guidance on how health workers can spend time with patients safely.

The description of a distressed mother as a typical “COVID patient” shows how placing patients in categories, a way of managing heavy workloads, can slide into ridicule. It also points to a wider risk. If calm, undemanding patients fare better, as one participant suggested, the most distressed patients, who need most attention, may receive least. Similar dynamics have been reported in maternity care, where women perceived as difficult or uncooperative were more likely to be mistreated [13]. Conversely, the health worker who explained that “we are just trying to contain the spread of the thing” shows how a single sentence can separate the patient from the disease. Since many patients with COVID-19 feared stigma [20], and health workers themselves reported stigma within isolation centres [9], the way isolation is explained to patients is part of preventing stigma rather than a matter of courtesy.

Lastly, these findings have consequences beyond individual distress. The first concerns patient safety. Isolated patients experience more adverse events linked to failures of supportive care [1], and participants in this study described breathless patients waiting for help. The second concerns psychological harm. Isolation is associated with higher anxiety and depression [2], and visiting restrictions with loneliness and reduced wellbeing [4]; the conduct of health workers can either soften or deepen these effects. The third concerns trust.

Everyday encounters with health workers shape trust in the health system [18], and poor treatment can lead patients to avoid facilities [17]. Patients who experienced isolation as neglect or ridicule, and who share these experiences with others, may be less willing to seek testing, disclose symptoms or accept isolation in a future outbreak. With zoonotic outbreaks in Africa becoming more frequent [19], a response that repeats these patterns could undermine the public cooperation on which outbreak control depends.

Supportive care, by contrast, was often brief: noticing, asking, offering a time frame, connecting recovery to family. Isolated patients in other contexts have reported such short gestures, such as a gaze, words of encouragement or a prompt response [5,6], and associated caring staff with feeling calm [20]. Improving relational care in isolation may therefore depend less on lengthening contact than on making each contact count, and on making such care a routine expectation rather than a matter of individual discretion.

## Limitations of the Study

This study centres the accounts of patients, whose perspectives on health workers’ conduct during COVID-19 have rarely been the focus of research, in an African setting where such evidence might be insufficient. Applying an established typology allowed comparison of findings with a substantial body of research and to identify a domain that the typology does not currently capture.

However, I acknowledge the following limitations. The interviews took place in early 2023, so accounts may have been shaped by recall and reflection. Participants were recruited in one city, from selected hospitals, and partly through snowball sampling, which may have favoured people with particular experiences or networks. The requirement that participants communicate in English may have excluded some patients. We interviewed patients only, so our interpretation of health workers’ motives rests on patients’ accounts. Future research could test fear-driven distancing as a domain of mistreatment in other outbreaks, such as Ebola, Marburg and mpox, and should study patients and health workers in the same isolation units to examine how working conditions shape conduct.

## Conclusion

Isolation asks patients to endure illness without the people who would normally care for them and leaves health workers as their only human contact. For participants isolated with COVID-19 in Nairobi, that contact was often brief, confined to routine monitoring and, at times, dismissive. These patterns were shaped by the conditions of isolation, including the threat of infection, scarce staff and protective equipment, and protocols that limited contact, as much as by individual conduct. Where health workers noticed, asked and encouraged, patients found hope, but such care depended on who was on duty.

This study shows that the typology of mistreatment developed for maternity care applies to care in isolation, and it proposes fear-driven distancing as an additional domain for outbreak settings. Left unaddressed, these patterns risk patient safety, deepen psychological harm and may erode the trust on which future outbreak responses depend. As outbreaks in Africa and beyond become more frequent, preparedness should treat relational care as part of infection control by setting minimum standards for contact, making psychosocial assessment routine, preparing staff to explain isolation without stigma, and protecting health workers from the fear that draws them away from patients.

## Data Availability

The data underlying this study are interview transcripts from patients who were isolated with COVID-19. They cannot be shared publicly because they contain potentially identifying information about participants and the hospitals where they were treated, and participants did not consent to public release of their transcripts. Anonymised quotations supporting the findings are included in the manuscript. Anonymised transcripts are available on reasonable request from the Daystar University Institutional Scientific and Ethical Review Committee, for researchers who meet the criteria for access to confidential data

## Acknowledgments

I deeply appreciate the support and mentorship I received from my university supervisors (Dr. Kinya Mwithia and Prof Wilson Ugangu) at the time of conducting the study. In addition, I appreciate the recovered patients who openly shared their experiences and made it possible to complete this study. Also, I am grateful to the two research assistants (Joy Kavata and Luciah Nzisa) who supported me in conducting interviews and doing transcriptions. Lastly, and most significantly, I appreciate the support received during the writing of this manuscript from my current mentors at Strathmore PRISM Centre and Initiative to Develop African Research Leaders (IDeAL).

